# Detailed mapping of mesothelioma cases in Denmark to identify areas with elevated risk: a nationwide population-based study

**DOI:** 10.1101/2025.01.20.25320820

**Authors:** Heidi Søgaard Christensen, Rikke Hedegaard Jensen, Lars Hernández Nielsen, Lise Dueholm Bertelsen, Christian Teglgaard, Jakob Bønløkke, Marianne Tang Severinsen, Martin Bøgsted

## Abstract

**Objectives:** Previous studies mapping pleural mesothelioma in Denmark have found that the risk varies between Danish regions. However, evaluating disease risk for such relatively large geographical units ignores any heterogeneity within the unit and can thus diminish more local spikes in risk, missing smaller areas of excess risk. In this study, we examined the distribution of pleural mesothelioma in Denmark on an unprecedented detailed scale, mapping cases to each of the Danish parishes.

**Methods:** We identified individuals diagnosed with pleural mesothelioma between 1990 and 2021 in the Danish Cancer Registry. Considering age- and sex-standardised incidence rate ratios, we used a conditional autoregressive random effects model to smooth incidence rate ratios across parishes. Parishes with a smoothed parish-to-national incidence rate ratio higher than 1.25 or 2.0 with a posterior probability of more than 95% were flagged as parishes with an excess risk of pleural mesothelioma.

**Results:** We identified 3105 incident cases of pleural mesothelioma in the study period. A total of 73 and 14 parishes were flagged with incidence rate ratios significantly above 1.25 and 2.0, respectively. The flagged parishes had smoothed incidence rate ratios ranging from 1.80 to 4.65.

**Conclusions:** We provided a detailed mapping of pleural mesothelioma cases in Denmark and found five distinct areas, each covering several parishes, with a significantly elevated risk. All these areas were in the proximity of previous asbestos-using industries.

## Introduction

Mesothelioma is a rare malignancy with a very high mortality rate that is most commonly found in the pleura(Walker-Bone et al., 2023). At least 80% of pleural mesothelioma cases can be linked to prior exposure to asbestos(Marinaccio et al., 2012; Walker-Bone et al., 2023). Studies mapping mesothelioma cases have identified an elevated risk in areas of previous asbestos-using industries such as asbestos cement manufacturing plants, shipyards, asbestos mines, and asbestos textile industry(Cameron et al., 2022; Neyens et al., 2017; Corfiati et al., 2015; Panou et al., 2019). In Denmark, reported regional differences in incidence rates of mesothelioma are presumably associated with the location of previous asbestos-using industries(Andersson and Olsen, 1985; Skammeritz et al., 2013; Hemminki et al., 2021). However, Danish regions are relatively large geographical units, containing between 10% and 30% of the Danish population, and as any heterogeneity of risk within a region is ignored, the risk for smaller hotspot areas within a region is diluted. Considering a more granular mapping, Panou et al. investigated the distribution of mesothelioma across the Danish parishes, but only for women in the North Denmark Region and focused on distinguishing environmentally caused cases from occupational cases (Panou et al., 2019).

In this paper, we present an unprecedented detailed mapping of all incident pleural mesothelioma cases in Denmark from 1990 to 2021, considering incidence rates for each of the 2141 Danish parishes.

## Methods

### Danish mesothelioma data

All Danish residents diagnosed with pleural mesothelioma between 1 January 1990 and 31 December 2021 were identified using the Danish Cancer Registry which contains information on all cancer cases in the Danish population since 1943(Gjerstorff, 2011). The diagnoses available in the Danish Cancer Registry are coded according to the International Classification of Disease 10th Revision (ICD-10) or have been converted to ICD-10 if given before its introduction, and the morphology is coded according to the International Classification of Diseases for Oncology 3rd Revision (ICD-O-3)(Gjerstorff, 2011). We defined pleural mesothelioma as individuals diagnosed with either ICD-10 code C45.0 or C38.4 with ICD-O-3 morphology codes indicating mesothelioma (M9050/3, M9051/3, M9052/3, or M9053/3). Using the Danish Address Register, individuals were assigned to one of 2141 Danish parishes based on their residential address at the date of diagnosis. We considered the parish division as of 2021. The parish populations range from 31 to 46,586 individuals with a median population of 1039 individuals.

This study is a register study that does not require informed consent in Denmark. The study has, according to the General Data Protection Regulation (GDPR), been registered in the North Denmark Region’s list over register studies (reg. no. F2023-095).

### Statistical analysis

To evaluate whether a parish had an elevated risk of pleural mesothelioma compared to the national incidence rate, we calculated the parish-to-national incidence rate ratio (IRR), corresponding to the ratio of the observed number of cases in a parish with the number of cases expected assuming the national incidence rate. The expected number of cases was indirectly standardised with respect to age and sex using seven age groups (≤ 40, 41 − 50, 51 − 60, 61 − 70, 71 − 80, 81 − 90, and > 90 years). As the number of pleural mesothelioma cases per parish is low, the raw incidence rate estimates will be unstable. Therefore, we considered a model framework that allowed us to improve estimates of the IRRs by borrowing information from neighbouring parishes and thus smoothing estimates across parishes. Specifically, using a fully Bayesian setup, we modelled the number of pleural mesothelioma cases by a spatial Poisson distributed generalised linear mixed model with a conditional autoregressive (CAR) correlation structure specified by the Leorux model(Leroux et al., 2000); details can be found in Appendix A. For each parish, we calculated the posterior probability of the smoothed IRR being higher than 1.25 and 2, corresponding to incidence rates elevated with more than 25% or 100% compared to the national rate, respectively. Parishes for which the posterior probability was 95% or higher were flagged as areas with elevated risk of pleural mesothelioma at the specified level. The residual autocorrelation was evaluated using Moran’s I statistic.

All analyses were performed in R (v4.2.2;R Core Team, 2021). The R-package CARBayes (v6.1.1;Lee, 2013) was used to fit the spatial model and the R-package ape to calculate Moran’s I statistic (v5.8;Paradis and Schliep, 2019).

## Results

A total of 3105 Danish residents were diagnosed with pleural mesothelioma between 1 January 1990 and 31 December 2021, which yields a national incidence rate of 1.72 per 100,000 person-years. Of these, 2618 (84%) were male and the overall median age at diagnosis was 70 years. Across age and sex groups, the national incidence rate was highest among individuals aged 70 to 90 years for both males and females, and incidence rates were substantially higher in males than females across all age groups (see Table 1). The raw age- and sex-standardised parish-to-national IRRs are shown in Figure B.1 in Appendix B, and the smoothed IRRs obtained from the fitted CAR autocorrelation model in Figure 1. No residual autocorrelation was detected (Moran’s I statistic = -0.016, p-value = 0.28).

**Table 1:** The national incidence rate per 100,000 person-years of pleural mesothelioma for each age and sex group.

| Age group (years) | Male incidence rate | Female incidence rate |
| --- | --- | --- |
| $\leq 40$ | 0.02 | 0.02 |
| 41 – 50 | 0.63 | 0.16 |
| 51 – 60 | 3.21 | 0.61 |
| 61 – 70 | 7.85 | 1.29 |
| 71 – 80 | 16.43 | 2.21 |
| 81 – 90 | 14.64 | 2.40 |
| $> 90$ | 3.52 | 1.04 |

**Figure 1:**
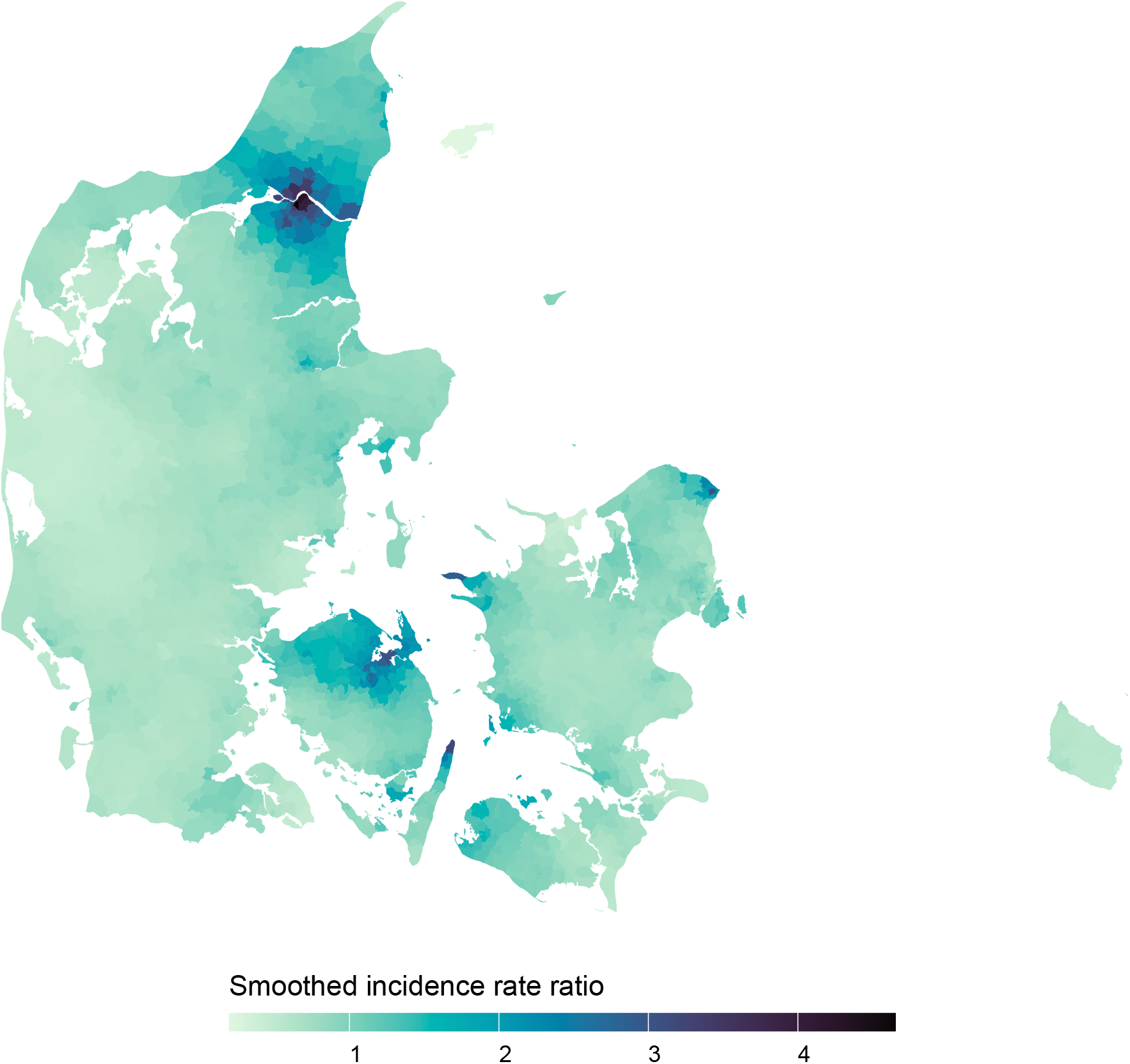
Posterior mean of the smoothed parish-to-national incidence rate ratios of pleural mesothelioma in Denmark for each parish.

We found 73 and 14 parishes with smoothed incidence rates more than 25% and 100% higher than the national, respectively, with a posterior probability of more than 95% (see Figure 2, and Figure B.2 and Table B.1 in Appendix B). The parishes flagged with a more than 100% elevated incidence rate had posterior mean smoothed IRRs ranging from 2.78 to 4.65 and were located in three distinct areas of Denmark around the three cities Aalborg, Odense, and Helsingør. Considering an elevated risk of more than 25%, more parishes in these three areas were flagged as well as one parish near Kalundborg and one in Frederikshavn; the posterior mean smoothed IRRs ranged from 1.80 to 4.65.

**Figure 2:**
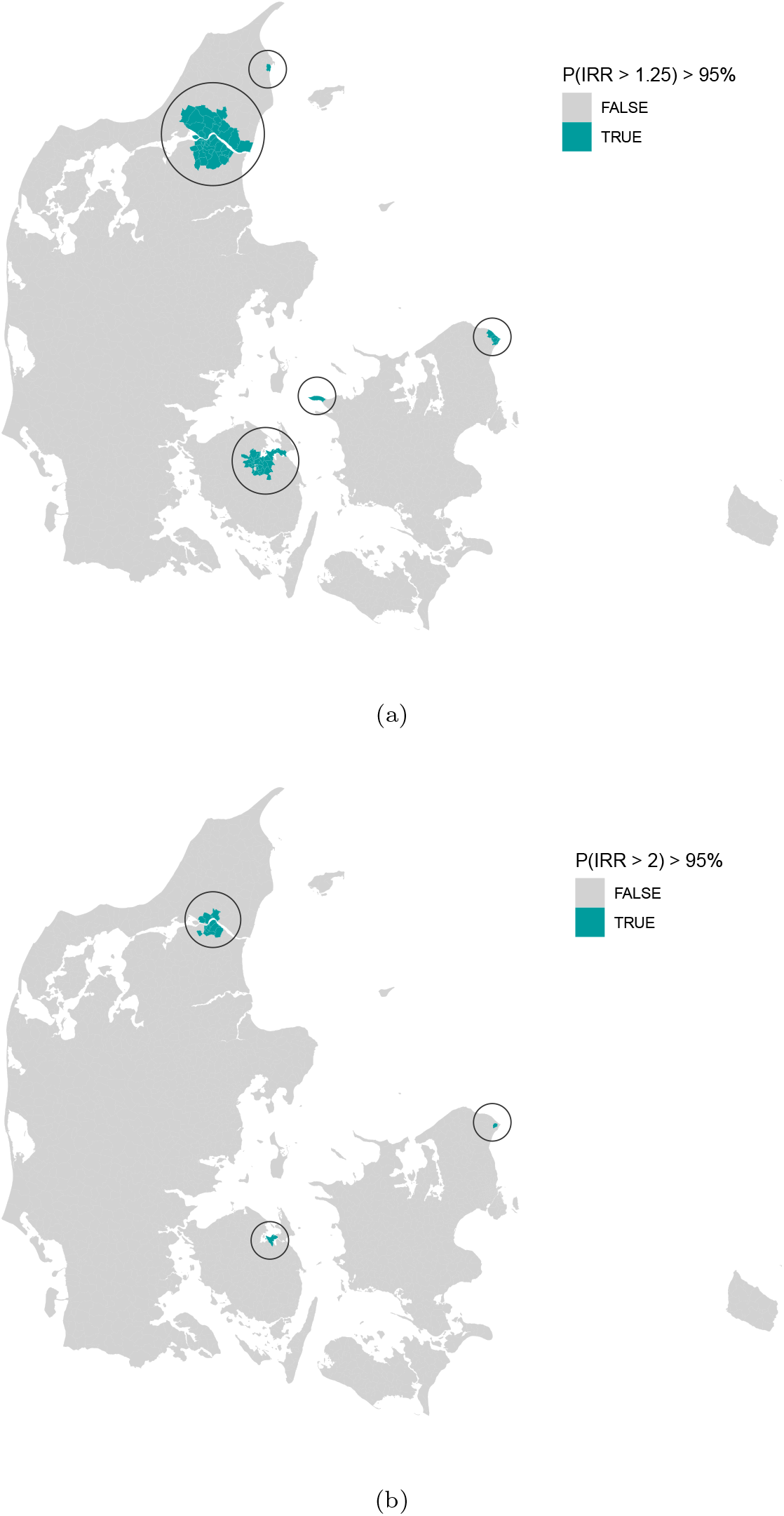
Parishes with smoothed incidence rates more than 25% (a) and 100% (b) higher than the national level with a posterior probability of more than 95%. Abbreviations: IRR, incidence rate ratio.

At diagnosis, individuals in our cohort had lived a median time of 27 years in the same parish.

## Discussion

We found five distinct areas in Denmark that with high certainty had an elevated risk of pleural mesothelioma. Specifically, 14 parishes were flagged with smoothed incidence rates more than twice as high as the national level, while 73 parishes were flagged with an incidence rate elevated with more than 25%. The 14 parishes were located around and in the cities of Aalborg, Odense, and Helsingør, while the 73 parishes further covered a parish in Frederikshavn and a parish near Kalundborg. All of these cities previously had shipyards, and Aalborg further had a large asbestos cement factory, Odense a large producer of asbestos-containing brakes, and Kalundborg an oil refinery. Workers in such industries are some of the occupational groups with increased risk of mesothelioma (see e.g.Sanden et al., 1992; Ulvestad et al., 2002; Sorahan, 2007).

In agreement with our results, previous studies have reported increased standardised incidence ratios for male inhabitants in several Danish cities with previous shipyards and/or an asbestos cement factory, with the highest ratios in Aalborg, Helsingør, and Odense(Andersson and Olsen, 1985; Kjærgaard and Michelsen, 1997). With our choices of thresholds, no parishes were flagged in the capital area of Copenhagen, although existing studies, considering earlier time periods, have reported an increased risk(Andersson and Olsen, 1985; Kjærgaard and Michelsen, 1997; Skammeritz et al., 2013). As noted by Skammeritz et al. (2013), the male incidence rate of mesothelioma in the Capital Region of Denmark seems to have stagnated since the 1980s, while the incidences in the North Denmark Region and the South Denmark Region, that contain the cities Aalborg and Odense, respectively, have continued to increase up until 2009 where their study period ended. Since we examined the time period 1 January 1990 to 31 December 2021, this decreased incidence rate of mesothelioma in the Capital Region was confirmed by our study.

Asbestos exposure in individuals with mesothelioma has been thoroughly examined in the area of Aalborg(Raffn et al., 1989; Skammeritz et al., 2011; Panou et al., 2019; Dalsgaard et al., 2019). Among 122 patients seen at an occupational clinic in Aalborg, 87.7% had a known occupational exposure(Skammeritz et al., 2011). For female patients in the North Denmark Region, a total of 75% had an identified asbestos exposure with non-occupational exposure accounting for the majority of cases (Panou et al., 2019). Additionally, increased risks of mesothelioma have been reported in children who attended schools and lived in the neighbourhood of the asbestos cement factory in Aalborg(Dalsgaard et al., 2019). Asbestos use has, with few exceptions, been prohibited in Denmark since 1986, though the use of asbestos in brakes were not prohibited until 2004 (Fonseca et al., 2022).

In this study, the mapping was based on the residential location at time of diagnosis. However, due to the long latency period from first asbestos exposure to development of pleural mesothelioma with a median of 38 years, the distribution at time of diagnosis does not necessarily reflect areas of asbestos exposure(Reid et al., 2014). This is evident in the study by Cameron et al. (2022), where high counts of cases in Australia were seen in popular retirement areas in Perth while only few cases were reported in previous mining and milling areas that have been depopulated since the industry closed down. At diagnosis, individuals in our cohort had lived a median time of 27 years in the same parish. Note, other choices of baseline incidence rate than the national may have led to different results. Similarly, a less restrictive probability threshold would lead to more parishes flagged with elevated risk.

A strength of our study is the validated data on diagnosis and parish of residence which have been retrieved from high-quality population-based registries. Furthermore, since the Danish healthcare system provides free access to health services, the risk of socioeconomic factors affecting the findings is considered low. Further, no existing study reports the risk of pleural mesothelioma at such granular level for all of Denmark.

In conclusion, from a granular mapping of pleural mesothelioma cases in Denmark, we found five distinct areas with elevated risk; all these areas could be linked to previous asbestos-using industries.

## Data Availability

Data was made available through Statistics Denmark. Research data from Statistics Denmark cannot be shared according to the GDPR.

## Acknowledgements

This work was supported by the Danish National Research Foundation [grant number DNRF148], Health insurance Denmark [grant number 2022-0276], a scholarship from the Novo Nordisk Foundation [NNF21SA0069373], and a scholarship from the Svend Andersen Foundation.

