## Appendices for "Detailed mapping of mesothelioma cases in Denmark to identify areas with elevated risk: a nationwide population-based study"

### Appendix A. Workflow for identifying parishes of elevated risk

*Standardised expected number of cases*

To define which parishes have a significantly elevated risk of pleural mesothelioma, we consider the parish-to-national incidence rate ratio (IRR); that is, the ratio of the observed number of cases in a parish and the number of cases expected under the national incidence rate. As the incidence of pleural mesothelioma depends highly on age and sex, we consider the indirectly age- and sex-standardised expected number of cases. That is, for the $i$th parish we calculate the standardised expected number of cases by

$$e_i = \sum_j t_{ij} r_j,$$

where $r_j$ is the national incidence rate for age and sex group $j$ and $t_{ij}$ is a measure of person-time in parish $i$ and age and sex group $j$. We estimated person-years by $t_{ij} = l \cdot n_{ij}$, where $l$ is the length of the study period in years and $n_{ij}$ is the 2014 population in parish $i$ for age and sex group $j$ (Danmarks Statistik, 2023).

Note, due to a small number (or a zero-count) of age- and sex-specific observed cases of pleural mesothelioma in some parishes, we used indirect standardisation instead of direct standardisation which may be preferred if the counts are high enough (or at least non-zero) (Chan et al., 1988).

*Spatial model*

We considered a fully Bayesian model framework that allowed us to improve estimates of the IRRs by borrowing information from neighbouring parishes and thus smoothing estimates across parishes. Specifically, we considered a generalised linear mixed model with Poisson distributed response and a conditional autoregressive (CAR) prior proposed by Leroux et al. (2000) for modelling spatial autocorrelation. In detail, the model was given by

$$\begin{aligned}
y_i|\lambda_i &\sim \text{Poisson}(\lambda_i),\\
\log(\lambda_i) &= \alpha + \log(e_i) + u_i,\\
u_i|u_{-i} &\sim N\left(\frac{\rho \sum_k w_{ik} u_k}{\rho \sum_k w_{ik} + 1 - \rho}, \frac{\tau^2}{\rho \sum_k w_{ik} + 1 - \rho}\right),\\
\alpha &\sim N(\mu_\alpha, \sigma_\alpha),\\
\tau^2 &\sim \text{Inverse-Gamma}(a, b),\\
\rho &\sim \text{Uniform}(0, 1),
\end{aligned}$$

where $y = (y_1, \ldots, y_m)$ denotes the number of cases across the $m = 2141$ parishes, $u_i$ a spatially autocorrelated random effect for parish $i$, $u_{-i}$ all random effects except the $i$th, and $W = [w_{ik}]$ a neighbourhood matrix. We specified $W$ as a binary matrix with $w_{ik} = 1$ if parish $i$ and $k$ shared a border or was connected by bridge, and, for parishes constituting a whole island, connections by ferry also qualified as a neighbour relation. Otherwise, $w_{ik} = 0$. The neighbour relations are illustrated in Figure A.1 The model was fitted using the R-package CARBayes and its default settings for priors with $\mu_\alpha = 0$, $\sigma_\alpha = 100,000$, $a = 1$, and $b = 0.01$ (Lee, 2013).

*Flagging parishes with elevated risk*

For a user-specified threshold $\theta > 0$, we consider incidence rates more than $100 \cdot \theta\%$ above the national level to be elevated, and for the $i$th parish, we define its potential as an area of excess risk at level $\theta$ by

$$p_i(\theta) = P(\lambda_i/e_i > 1 + \theta \,|\, y),$$

the one-sided Bayesian credible interval of the smoothed IRR, $\lambda_i/e_i$. We flag the $i$th parish as having elevated risk at level $\theta$ whenever the posterior probability $p_i(\theta) > \tau$, where $0 < \tau \leq 1$ is a user-specified threshold. In this study, we consider $\theta \in \{0.25, 1\}$ and $\tau = 0.95$.

Inference was based on Markov Chain Monte Carlo (MCMC) simulations from three chains, using a burnin of 100,000 samples, thinning of 20, and a total number of samples used for inference of 6,000 samples. For each parish $i$, we evaluated the posterior mean of the smoothed IRR $\lambda_i/e_i$ and 95% credible intervals with limits given by the 2.5th and 97.5th percentiles. Further, we calculated the posterior probability $p_i(\theta)$, i.e., the proportion of MCMC iterations with $\lambda_i/e_i > 1 + \theta$.

*Generalisability and code accessibility*

The workflow described above to identify parishes with elevated mesothelioma risk was chosen to handle the low count of cases per parish and the smaller population in some parishes, leading to unstable incidence rate estimates. The method can be generalised to other diseases and geographical units. Due to the GDPR, we are not permitted to publish the pleural mesothelioma data. However, to provide an example of the workflow our analysis is based on, R-code for analysing the distribution of males and females in Denmark has been made publicly available on Github: `https://github.com/CLINDA-AAU/disease_CAR_example`.

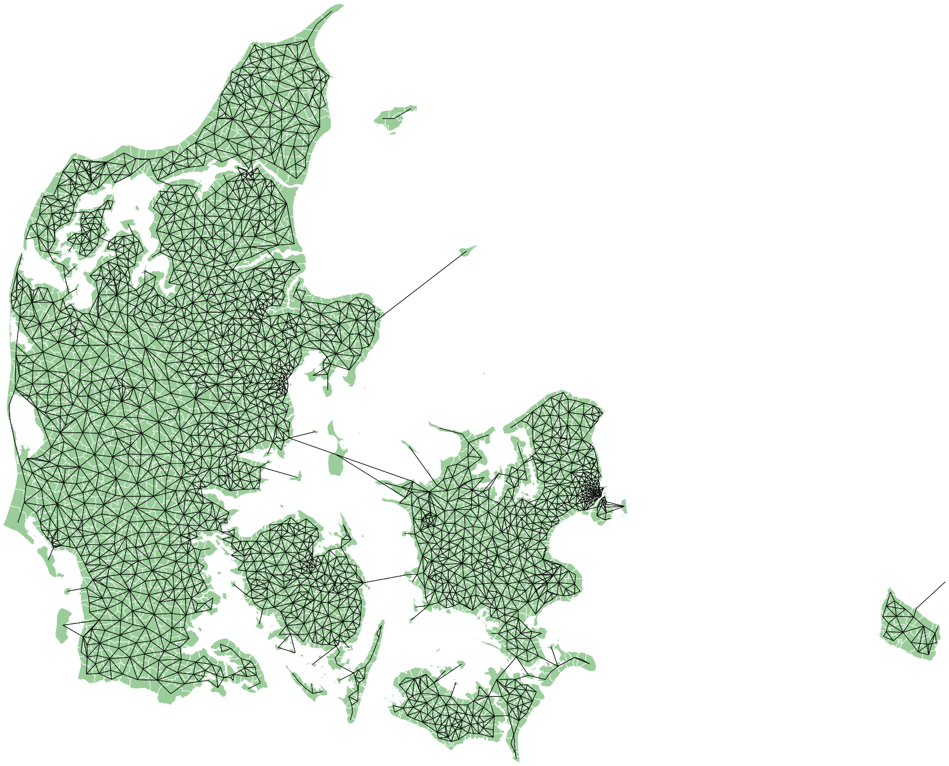

Figure A.1: Illustration of the neighbour relations. Parishes connected by a line are considered neighbours.

### Appendix B. Results

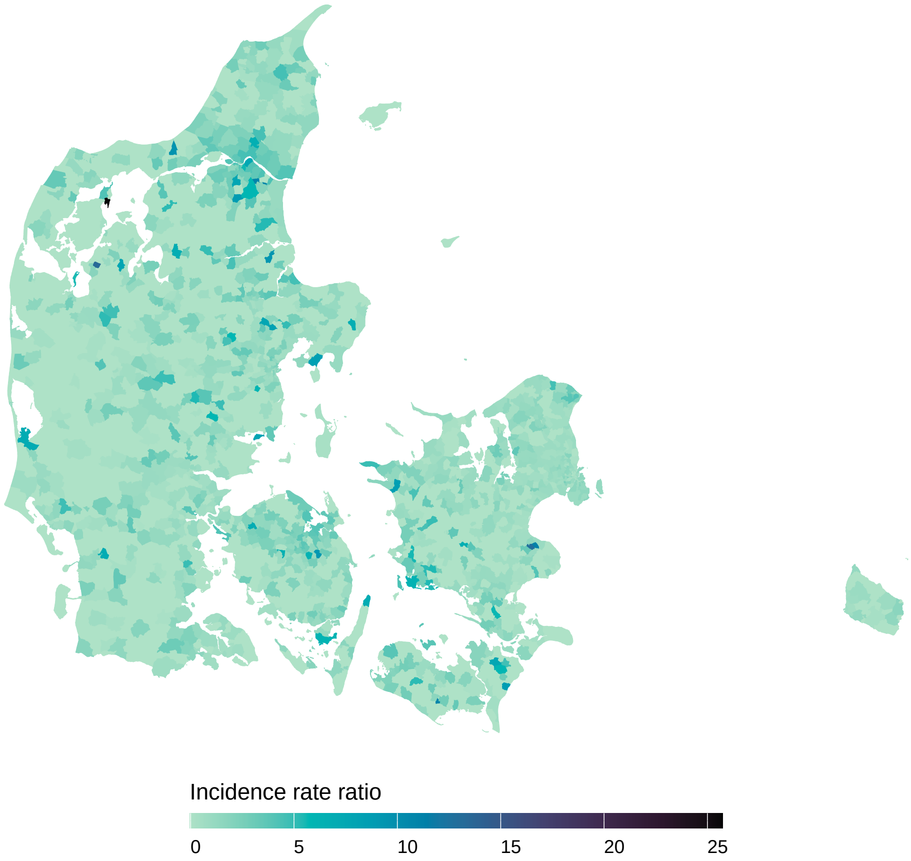


Figure B.1: For each parish, the raw age- and sex-standardised incidence rate ratios (parish-to-national) of pleural mesothelioma.

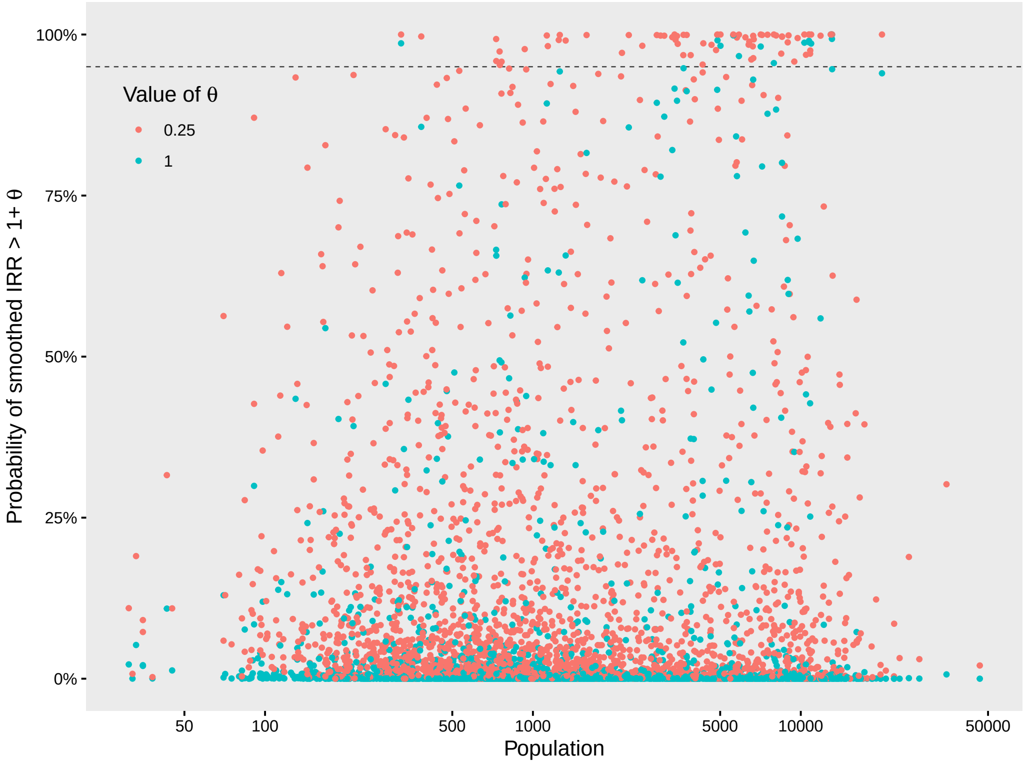


Figure B.2: For each parish, the posterior probability that the smoothed incidence rate is more than 25% (red) or 100% (blue) higher than the national rate against the population size; the dashed line marks 95%.

| **Municipality** | **Parish** | **Smoothed IRR (95% CI)** |
|---|---|---|
| Aalborg | Ajstrup | 1.93 (1.23, 2.83) |
| Aalborg | Ellidshøj | 2.05 (1.24, 3.20) |
| Aalborg | Hasseris | 2.08 (1.45, 2.9) |
| Aalborg | Øster Hassing-Gåser | 2.23 (1.31, 3.56) |
| Aalborg | Ferslev | 2.24 (1.27, 3.65) |
| Aalborg | Gudumholm | 2.25 (1.30, 3.60) |
| Aalborg | Horsens | 2.27 (1.36, 3.55) |
| Aalborg | Frejlev | 2.32 (1.39, 3.62) |
| Aalborg | Vadum | 2.40 (1.53, 3.56) |
| Aalborg | Sejlflod | 2.46 (1.17, 4.70) |

| | | |
|---|---|---|
| Aalborg | Gunderup | 2.46 (1.61, 3.60) |
| Aalborg | Ansgars | 2.49 (1.52, 3.73) |
| Aalborg | Nøvling | 2.56 (1.64, 3.78) |
| Aalborg | Svenstrup | 2.56 (1.60, 3.82) |
| Aalborg | Vester Hassing | 2.61 (1.52, 4.15) |
| Aalborg | Budolfi | 2.65 (1.66, 3.94) |
| Aalborg | Sulsted | 2.71 (1.77, 3.98) |
| Aalborg | Nørresundby | 2.72 (1.67, 4.14) |
| Aalborg | Margrethe | 2.74 (1.75, 4.02) |
| Aalborg | Storvorde | 2.78 (1.62, 4.43) |
| Aalborg | **Skalborg** | **2.78 (1.92, 3.88)** |
| Aalborg | Lillevorde | 2.84 (1.54, 4.81) |
| Aalborg | Hammer | 2.84 (1.66, 4.57) |
| Aalborg | Gistrup | 2.84 (1.72, 4.33) |
| Aalborg | Hals | 2.87 (1.66, 4.56) |
| Aalborg | **Sønder Tranders** | **3.03 (2.18, 4.05)** |
| Aalborg | Dall | 3.09 (1.81, 4.95) |
| Aalborg | **Romdrup-Klarup** | **3.12 (2.06, 4.56)** |
| Aalborg | **Vor Frue** | **3.17 (2.07, 4.71)** |
| Aalborg | Vesterkær | 3.19 (1.82, 5.07) |
| Aalborg | **Lindholm** | **3.30 (2.14, 4.75)** |
| Aalborg | **Hans Egedes** | **3.33 (2.18, 4.76)** |
| Aalborg | Vor Frelsers | 3.34 (1.68, 5.69) |
| Aalborg | **Vodskov** | **3.36 (2.17, 4.90)** |
| Aalborg | **Sankt Markus** | **3.46 (2.16, 5.13)** |
| Aalborg | **Hvorup** | **3.59 (2.44, 5.04)** |
| Aalborg | **Nørre Tranders** | **3.96 (2.70, 5.49)** |
| Aalborg | **Vejgård** | **4.65 (3.37, 6.20)** |
| Aalborg | **Rørdal** | **4.27 (2.19, 7.64)** |
| Brønderslev | Ørum | 1.92 (1.16, 2.98) |
| Frederikshavn | Bangsbostrand | 1.95 (1.20, 2.96) |
| Helsingør | Sthens | 1.94 (1.23, 2.87) |
| Helsingør | Sankt Mariæ | 2.13 (1.26, 3.33) |
| Helsingør | Hellebæk | 2.27 (1.46, 3.36) |
| Helsingør | Sankt Olai | 2.28 (1.42, 3.38) |
| Helsingør | **Vestervang** | **3.05 (2.08, 4.25)** |
| Kalundborg | Røsnæs | 2.87 (1.10, 5.79) |
| Kerteminde | Kerteminde-Drigstrup | 2.16 (1.30, 3.27) |

| | | |
|---|---|---|
| Kerteminde | Marslev | 2.22 (1.37, 3.40) |
| Kerteminde | **Munkebo** | **3.01 (1.95, 4.43)** |
| Jammerbugt | Aaby | 2.01 (1.29, 2.96) |
| Jammerbugt | Biersted | 2.09 (1.21, 3.32) |
| Odense | Paarup | 1.80 (1.25, 2.49) |
| Odense | Fraugde | 1.83 (1.18, 2.70) |
| Odense | Ansgars | 1.84 (1.19, 2.68) |
| Odense | Sankt Knuds | 1.88 (1.19, 2.81) |
| Odense | Næsbyhoved-Broby | 1.91 (1.22, 2.88) |
| Odense | Bolbro | 1.96 (1.23, 2.91) |
| Odense | Vollsmose | 1.98 (1.23, 2.94) |
| Odense | Hans Tausens | 1.98 (1.31, 2.88) |
| Odense | Lumby | 2.03 (1.32, 2.95) |
| Odense | Næsby | 2.11 (1.35, 3.09) |
| Odense | Hjallese | 2.11 (1.50, 2.88) |
| Odense | Vor Frue | 2.20 (1.35, 3.40) |
| Odense | Stige | 2.21 (1.33, 3.43) |
| Odense | Dalum | 2.25 (1.38, 3.46) |
| Odense | Åsum | 2.26 (1.39, 3.44) |
| Odense | Munkebjerg | 2.30 (1.48, 3.29) |
| Odense | Tornbjerg | 2.43 (1.58, 3.58) |
| Odense | Seden | 2.46 (1.50, 3.74) |
| Odense | Fredens | 2.62 (1.87, 3.56) |
| Odense | Korsløkke | 2.71 (1.88, 3.75) |
| Odense | Agedrup | 2.81 (1.66, 4.41) |

Table B.1: The posterior mean of the smoothed parish-to-national IRR for the 73 parishes flagged with an excess risk of at least 25% with a 95% posterior probability threshold. Parishes that were also flagged considering elevated incidence rates of at least 100% are written in bold. Abbreviations: IRR, incidence rate ratio; CI credible interval
